# Identifying the regional substrates predictive of Alzheimer’s disease progression through a convolutional neural network model and occlusion

**DOI:** 10.1101/2022.01.27.22269954

**Authors:** Kichang Kwak, William Stanford, Eran Dayan, the Alzheimer’s Disease Neuroimaging Initiative

## Abstract

Progressive brain atrophy is a key neuropathological hallmark of Alzheimer’s disease (AD). However, atrophy patterns along the progression of AD are diffuse and variable. Consequently, identifying the major regional atrophy patterns underlying AD progression is challenging. In the current study, we propose a method that evaluates the degree to which specific regional atrophy are predictive of AD progression, while holding all other atrophy changes constant. We first trained a dense convolutional neural network model to differentiate individuals with mild cognitive impairment (MCI) who progress to AD vs. those with a stable MCI diagnosis. Then, we retested the model multiple times, each time occluding major regions from the model’s testing set’s input. This revealed that the hippocampus, fusiform, and inferior temporal gyri, were the strongest predictors of AD progression, in agreement with established staging models. These results shed light on the major regional patterns of atrophy predictive of AD progression.

## Introduction

Alzheimer’s disease (AD) is a complex progressive neurodegenerative disease, and the leading cause of dementia (McKhann et al., 1984; Jack et al., 2018). The phenotype of AD is characterized by brain atrophy (Frisoni et al., 2010), and progressive volume loss, as evident with structural magnetic resonance imaging (MRI) (Pini et al., 2016; Halliday, 2017; Dallaire-Théroux et al., 2017). The pattern of brain atrophy displayed by single individuals is highly complex and variable (Jack, Knopman, et al., 2010), generally starting in the medial temporal lobes (Bobinski et al., 1997; Jack et al., 1999a), and later progressing to neocortical regions (Fox et al., 2001; Whitwell et al., 2007). The rate of atrophy in AD is non-linear (Jack et al., 2013), with considerable variability observed in the spatial pattern of atrophy displayed by individuals (Noh et al., 2014). Thus, simple univariate measures of regional volume loss may not adequately capture and quantify the spatiotemporal complexities of progressive brain atrophy in AD.

To properly handle the nonlinearities in the spatiotemporal evolution of biomarkers along the AD continuum (Jack et al., 2013), many studies have employed machine (and deep) learning modeling solutions (Suk et al., 2014; Stamate et al., 2019; Z. Yang et al., 2021). In particular, studies utilizing convolutional neural network architecture, with its established capability of extracting complex feature representations from large datasets, have been employed to accurately predict progression from mild cognitive impairment (MCI) to AD (Y. Huang et al., 2019; Spasov et al., 2019; F. Li & Liu, 2019; H. Li et al., 2019). However, most previous studies focused on improving the predictive accuracy of models, rather than providing interpretable findings in clinical settings. Thus, the spatiotemporal patterns of brain atrophy that are predictive of progression from MCI to AD, tested in a model that can properly capture complex feature representations, remain largely unknown.

In the current study, we aimed to identify the major spatial patterns of brain atrophy underlying the progressive neurodegenerative cascade in AD. To that effect, we deployed a convolutional neural network model to differentiate progressive versus stable MCI based on whole brain gray matter (GM) density maps derived from structural MRI. We then performed occlusion analysis (Kwak, Niethammer, et al., 2021), retesting the model while removing single regions from its input, to estimate the contribution of regional cortical atrophy to the progression of AD. Finally, we studied the spatial pattern of atrophy in more localized substates (the medial temporal lobe) and evaluated our methods against a well-established staging scheme for AD progression (Braak & Braak, 1991).

## Materials and Methods

### Participant Characteristics

Data used in the preparation of this article were obtained from the Alzheimer’s Disease Neuroimaging Initiative (ADNI). The ADNI was launched in 2003 as a public-private partnership, led by Principal Investigator Michael W. Weiner, MD. The primary goal of ADNI has been to test whether serial MRI, other biological markers, and clinical and neuropsychological assessment can be combined to measure the progression of MCI and early AD. For up-to-date information, see www.adni-info.org. All subjects provided written informed consent and the study protocol was approved by the local Institutional Review Boards. We used 219 subjects from the ADNI database to train the deep learning model to differentiate between subjects with AD and cognitively normal CN control participants. In accordance with recent recommendations (Jack et al., 2018) all subjects with AD had abnormal levels of cerebrospinal fluid (CSF) Amyloid beta (Aβ) and CSF tau, (henceforth, A+T+), based on established cut-offs (Ewers et al., 2019), while all CN subjects displayed no signs of such pathology (henceforth, A−T−). The model was optimized during training based on 5-fold cross validation (Kwak, Giovanello, et al., 2021). We subsequently tested the model with an independent dataset composed of 115 A+ MCI subjects, obtained from the ADNI-2/GO cohort. This testing set was partitioned into subjects who showed progressive MCI (pMCI) or stable MCI (sMCI), depending on whether they progressed to A+T+ AD or not over an 18 months period. We excluded 13 additional subjects who reverted to normal cognition over the same time period. Demographic characteristics of each subsample used in this study are presented in Table 1.

**Table 1.**

|  | Training data |  | Test data |  |
| --- | --- | --- | --- | --- |
|  | AD (A+T+) | CN (A-T-) | pMCI | sMCI |
| N | 110 | 109 | 34 | 81 |
| Age | 72.9±8.0 | 71.0±5.3 | 72.3±5.1 | 72.8±6.9 |
| Gender, female | 46 (41.8%) | 57 (52.3%) | 14 (41.2%) | 24 (29.6%) |
| MMSE | 22.9±2.0 | 29.1±1.1 | 27.2±1.7 | 28.0±1.8 |

Continuous variables are presented as mean ± SD and categorical variable is presented as %.
Abbreviations: AD=Alzheimer's disease, CN=cognitively normal, pMCI=progressive mild cognitive impairment, sMCI=stable mild cognitive impairment, N=number of subjects, MMSE=Mini-Mental State Examination.

### Image acquisition

Structural MRIs were acquired in the ADNI study using 3T scanners and were based on either an inversion recovery-fast spoiled gradient recalled or a magnetization-prepared rapid gradient-echo sequences with sagittal slices and voxel size of 1.0×1.0×1.2 mm (Jack, Bernstein, et al., 2010). Full details of the T1 acquisition parameters and imaging processing steps are listed on the ADNI website (http://adni.loni.usc.edu/methods/documents/).

### Image processing

T1-weighted images were downloaded from the ADNI database. T1-weighted images were analyzed using Statistical Parametric Mapping 12 (SPM12; Wellcome Department of Imaging Neuroscience, Institute of Neurology, London, UK); http://www.fil.ion.ucl.ac.uk/spm) running on MATLAB 9.8.0 (Math-Works, Natick, MA, USA). Briefly, all MR images were aligned such that the origin was located at the anterior commissure and segmented into GM, white matter and CSF (Ashburner & Friston, 2005). We then used the diffeomorphic anatomical registration through exponentiated lie algebra (DARTEL) registration (Ashburner, 2007) to normalize the six tissue probability maps to Montreal Neurological Institute space, with a resolution of 2mm isotropic voxels, and produce the final GM density maps. Subsequently, all GM density images were used in the original intact models, as well as during occlusion analysis (See below). Cortical volumetric parcellation was performed as an initial step prior to occlusion analysis with the automated parcellation tool available in FreeSurfer v6.0 (https://surfer.nmr.mgh.harvard.edu/), based on the Desikan-Killiany protocol (Desikan et al., 2006).

### Procedure Overview

To delineate the contribution of regional atrophy patterns to AD progression, we extended an approach we have introduced recently in a study on the involvement of hippocampal subfields in AD progression (Kwak, Niethammer, et al., 2021), where a prognostic deep learning model is deployed in combination with occlusion learning. Our approach begins by training and testing the model in the task of differentiating pMCI vs. sMCI, based on whole-brain GM volume. The model is then retested multiple times, wherein specific brain regions are occluded from the model’s testing set’s input data, while all other model inputs remain unchanged. The performance of the models with occluded input are then compared to that of the intact, full model, and ranked according to differences with respect to the intact model.

### Deep learning model

Our model is based on the 3D Densely connected convolutional neural network (DenseNet) architecture (G. Huang et al., 2017), which has been designed such that all layers are directly connected to ensure maximum information flow in the network. The model’s proposed architecture is shown in Figure 1. The 3D volumes of GM density with size of 91 ×109×91 were used as input to the our model. Data augmentation was performed on the training set by flipping volumes left to right and randomly shifting by up to 10% and rotating by up to 20° in any direction. For each layer, the feature-maps resulting from of all preceding layers are used as inputs. The DenseNet model alleviates the problem of the vanishing gradient problem and has substantially less number of parameters than many other models (G. Huang et al., 2017). DenseNet consists of a convolutional layer, 4 dense blocks, 3 transition layers, a global averaging pooling layer and a fully-connected layer. Each dense block consists of several convolutional layers and a transition layer. Each dense block includes a composite function of multiple consecutive operations, including batch normalization layer, leaky rectified linear unit, a 1×1×1 convolutional layer, a 5×5×5 convolutional layer and a dropout layer. A transition layer between two dense blocks performs a down-sampling operation, which consists of batch normalization, 3×3×3 convolutional, leaky rectified linear unit, a dropout layer, and 2×2 ×2 average pooling layer. Global averaging pooling layers are then connected by a fully-connected layer. The last fully-connected layer generates a probability distribution over two labels with a sigmoid function.

**Figure 1.**
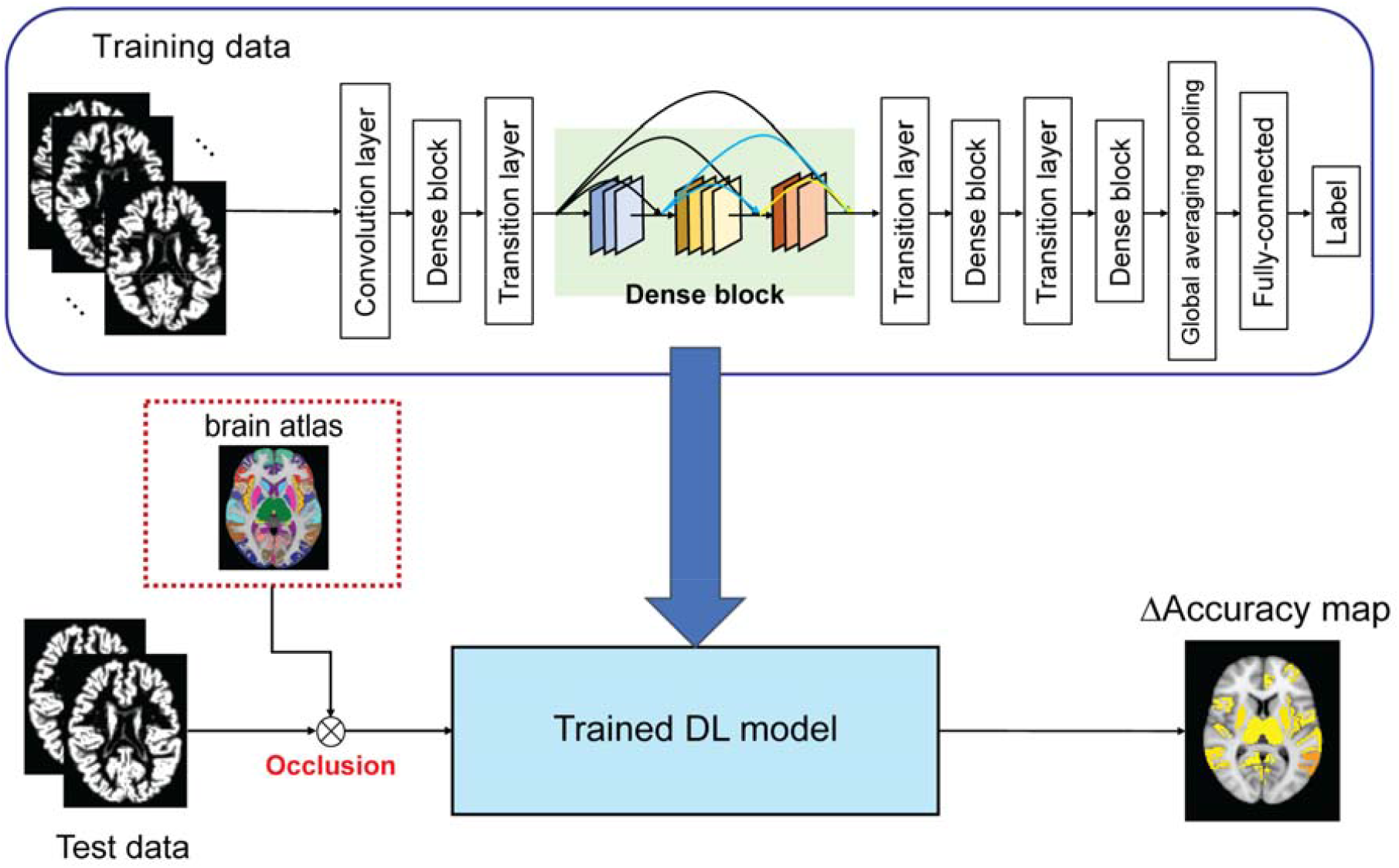
Study design. An illustration of the proposed deep learning model used in this study. A 3D-DenseNet convolutional neural network model was trained to differentiate AD and CN subjects based on whole brain GM density maps. The model was then tested in the task of classifying stable and progressive MCI. The trained model was subsequently re-tested multiple times in conjunction with occlusion analysis, wherein brain regions, defined based on whole brain atlas, were removed from the model’s input. The effect of occlusion on the performance of the model, relative to that of an intact model, was then calculated and visualized. Abbreviations: 3D-DenseNets=3D Densely convolutional neural network, AD=Alzheimer’s disease, CN=cognitively normal, GM=gray matter.

### Implementation

The deep learning model was implemented in the Keras library with the TensorFlow 2.0 backend. All training and testing were performed on an Ubuntu system 18.04.3, with 16GB RAM, Intel® Xeon® CPU @2.4GHz and 16GB Nvidia Tesla V100 graphic cards. The weights of our model were randomly initialized from a gaussian distribution. The model was trained for 200 epochs with a batch size of 24 and optimized using stochastic gradient descent based on adaptive estimation of first- and second-order moments (Kingma & Ba, 2015) and an exponentially decaying learning rate. The initial learning rate was set at 0.0001 and decayed by a factor of 0.9 after every 10000 steps. A dropout layer was included in the dense block, with the dropout rate set to 0.2. In the batch normalization step, beta and gamma weights were initialized with L2 regularization set at 1×10^−4^ and epsilon set to 1.1×10^−5^. An L2 regularization penalty coefficient, included in the fully-connected layer, was set at 0.01. The model was stable after an iteration of 150 epochs. Training time was about 10 hours.

### Occlusion analysis

Occlusion analysis was used as in our previous studies (Kwak, Niethammer, et al., 2021; Kwak, Giovanello, et al., 2021). Here, occlusion was used to identify the relative contribution of each brain region to prediction of AD progression, while holding all other complex, multivoxel atrophy patterns constant. Cortical regions were identified based on FreeSurfer’s cortical parcellation routine (see Imaging processing). To perform occlusion analysis on each single brain region, we masked it out from each input image by setting the values of all voxels corresponding to that brain region to zero. We then retested the model’s performance with occlusion and quantified the contribution of the occluded region to the model’s performance in predicting regional atrophy as the following:

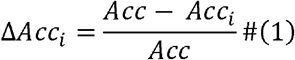

with *Acc* referring to the performance of the intact model with unmodified input images, and *Acc*_*i*_ referring to the model’s performance on input images with region *i* occluded.

We also implemented patch-based occlusion analysis to evaluate more specific regional contribution to the performance of the prediction model. We first defined the initialization mask around the structure of interest, e.g., the medial temporal lobe, merging ROIs (i.e., entorhinal, fusiform, parahippocampal, hippocampus, and amygdala) to define the initial mask. We then masked out 5×5×5 patches centered around each voxel of initial mask from input data. We then evaluated the Δ*Acc* for each occluded patch by retesting the trained model and assigning the Δ*Acc* to the central voxel of that patch. We iteratively performed this task for all voxels in the initial mask to generate a map depicting the contribution of voxels to prediction of AD progression within the target ROI.

### Contribution of Braak ROIs to prediction of AD progression

We next explored the contribution of atrophy in ROIs defined based on Braak’s staging scheme (Braak & Braak, 1991) to prediction of AD progression. This influential staging scheme work delineates neurofibrillary pathology in early, intermediate and late AD. We tested the extent to which atrophy in Braak ROIs contributed to the performance of our model in the task of differentiating pMCI vs. sMCI. In the current study, we focused on Braak stages I/II, which correspond to the transentorhinal stage, stages III and IV, which correspond to the limbic stage, and stages V and VI, which correspond to the isocortical stages. Braak ROIs were created by compositing FreeSurfer-derived ROIs (Table S1). We then evaluated via occlusion analysis the Δ*Acc* for each Braak ROI/stage, as explained in previous sections. Results are displayed on a cortical surface using *Simple Brain plot* (https://github.com/dutchconnectomelab/Simple-Brain-Plot).

## Results

### Participant Demographics

This study included data from a total of 334 subjects, across the AD, CN, pMCI and sMCI groups, all obtained from the ADNI database. Demographic characteristics are presented in Table 1. Based on the NIA-AA guidelines (Jack et al., 2018), only AD subjects with abnormal CSF Aβ and tau were included in the study, while CN subjects had to display no of signs of abnormality on the same biomarkers. In the comparison between the AD and CN groups, there were no significant differences in gender (*χ*^*2*^=2.41, *p*=0.12) while age (*t*_*217*_=2.19, *p*=0.04) was significantly different. All MCI subjects included in the analysis were amyloid positive (Jack et al., 2018). Comparison between the pMCI and sMCI groups, no significant differences in age (*t*_*113*_=0.32, *p*=0.75) or gender distributions (*χ*^*2*^=1.44, *p*=0.23).

### A deep learning model for differentiating between pMCI and sMCI

We first trained and a 3D convolutional neural network with the DenseNet architecture (G. Huang et al., 2017) for differentiating between pMCI and sMCI. We adopted a 5-fold cross validation framework in the training phase, in order to optimize the model and assess its stability, and applied data augmentation to the training dataset, as means to improve the performance of the model. As done in previous studies, we trained our model on the task of differentiating AD and CN subjects, and then tested the model’s performance in classifying pMCI and sMCI (Falahati et al., 2014; Y. Huang et al., 2019; F. Li & Liu, 2019; Kwak, Giovanello, et al., 2021). We reasoned that training the model on the former task will allow it to learn the necessary representations needed in order to successfully complete the latter task. For the task of differentiating AD and CN subjects, the model with the best performance achieved an accuracy of 93.75% in one of the folds and an area under the curve (AUC) of the receiver operating characteristic (ROC) curve of 0.98. In the task of classifying pMCI vs. sMCI, the proposed deep learning model achieved an accuracy of 73.90% and an AUC of 0.74 (Figure 2A). Accuracy may be misleading when used with an imbalanced dataset, therefore we also evaluated the normalized confusion matrix for the classifier’s performance (Figure 2B), finding that the model achieved sensitivity of 0.62 for correct prediction of pMCI and specificity of 0.78 for true negative prediction of pMCI (i.e., sMCI). We also evaluated the extent to which the performance of the proposed model differed from chance levels, with a permutation test under a null distribution generated with random classifier (*p*<0.001, Figure S1).

**Figure 2.**
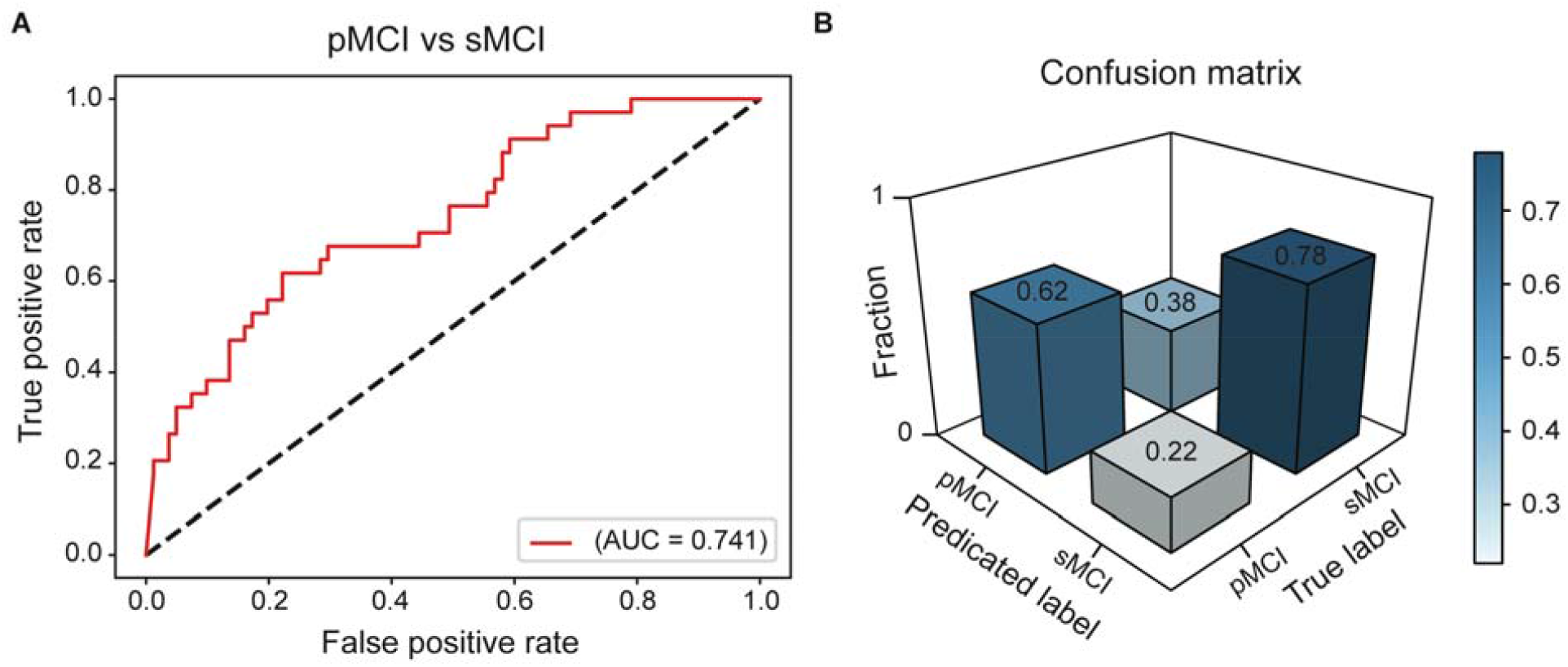
The performance of the proposed convolutional neural network model. The proposed convolutional neural network model was evaluated (**A**) ROC curve of the proposed model in the task of differentiating pMCI vs. sMCI. (**B**) Confusion matrix evaluating sensitivity (correct prediction of pMCI) and specificity (true negative prediction of pMCI, i.e., sMCI) in the task of differentiating pMCI vs. sMCI. Abbreviations: ROC=receiver operating characteristic, AUC=Area under the (ROC) curve, pMCI=progressive mild cognitive impairment, sMCI=stable mild cognitive impairment.

### Regional contribution to prediction of AD progression

We then used occlusion analysis to identify which regions contributed to the classification of pMCI vs sMCI. In this analysis, we repeatedly tested the deep learning model while occluding different brain regions in the test dataset to evaluate the individual contribution of each brain region to model performance, while all other atrophy patterns are held constant. To occlude a region, we set the intensity values of voxels in that region to zero before inputting the full image into to the deep learning model. The delta accuracies for each brain region are defined by the change in the model’s accuracy in classifying pMCI and sMCI after occlusion of a particular brain region normalized by the model’s baseline performance prior to occlusion (See Figure 3A-B). We found that the left and right hippocampus, fusiform gyrus, inferior temporal gyrus, and precuneus had the largest influence on the performance of the model, and by proxy to prediction of progression of MCI to AD. We tested if the size of the occluded ROIs related to accuracy in our proposed framework. No significant correlation was observed between the size of individual occluded ROI (Figure S2A) and accuracy loss (*R*^*2*^=0.01, *p*=0.36; Figure S2B). To further validate our proposed method, we next assessed the more precise and localized contribution of voxels within the medial temporal lobe to the prediction of AD progression using patch-based occlusion analysis. Briefly, we retested the trained deep learning model, each time occluding a patch centered on each voxel of the medial temporal region and calculating differences in accuracy relative to the intact model (Figure 3C). This analysis showed that within the medial temporal region, the bilateral hippocampus and amygdala are more central to the prediction of AD progression (Figure 3D), validating the spatial precision of our proposed method.

**Figure 3.**
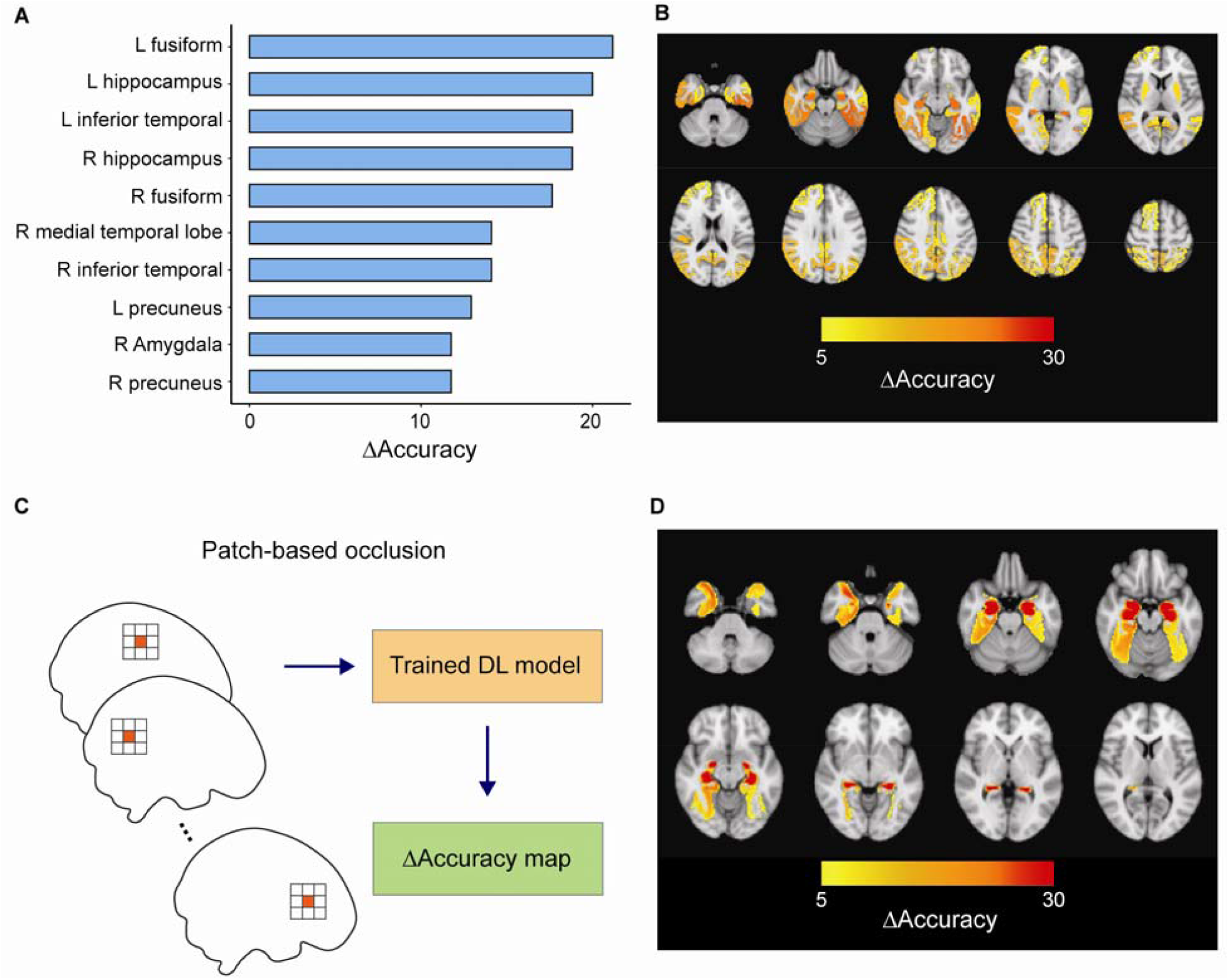
Occlusion analysis. (**A**) Delta accuracy (performance when regional-based occlusion is applied relative to that of an intact model) is shown for the top 10 regions with the highest contribution (highest accuracy loss) to prediction of AD progression. (**B**) Occlusion maps depicting the results of regional occlusion analysis overlayed on top of an anatomical image. (**C**) An illustration of patch-based occlusion analysis. At each step a single patch is masked out and then delta accuracy is mapped to the center voxel of that patch. (**D**) The results of patch-based occlusion analysis performed in the medial temporal lobe. Abbreviations: pMCI=progressive mild cognitive impairment, sMCI=stable mild cognitive impairment.

### Regional occlusion of ROIs based on the Braak staging scheme

Finally, we validated our occlusion analysis approach by testing whether occlusion of ROIs which are defined based on Braak’s staging scheme (Braak & Braak, 1991) results in appropriate model loss across the different ROIs (Figure 4). We found that the occlusion of the ROI corresponding to the intermediate Braak stage III, which primarily consists of limbic areas, had the largest impact on the performance of the model, relative to other staging-related ROIs (Figure 4). However, we also found that occlusion of transentorhinal ROIs defined in Braak stages I/II and limbic ROIs defined in Braak stages IV also highly impacted the performance. On the other hand, loss in accuracy was lower when occluding regions associated with the later Braak stages V and VI. Therefore, the relationships learned by our model aligned well with existing studies indicating the importance of regions defined in early Braak stages for predicting progression from MCI to AD (Braak & Braak, 1991).

**Figure 4.**
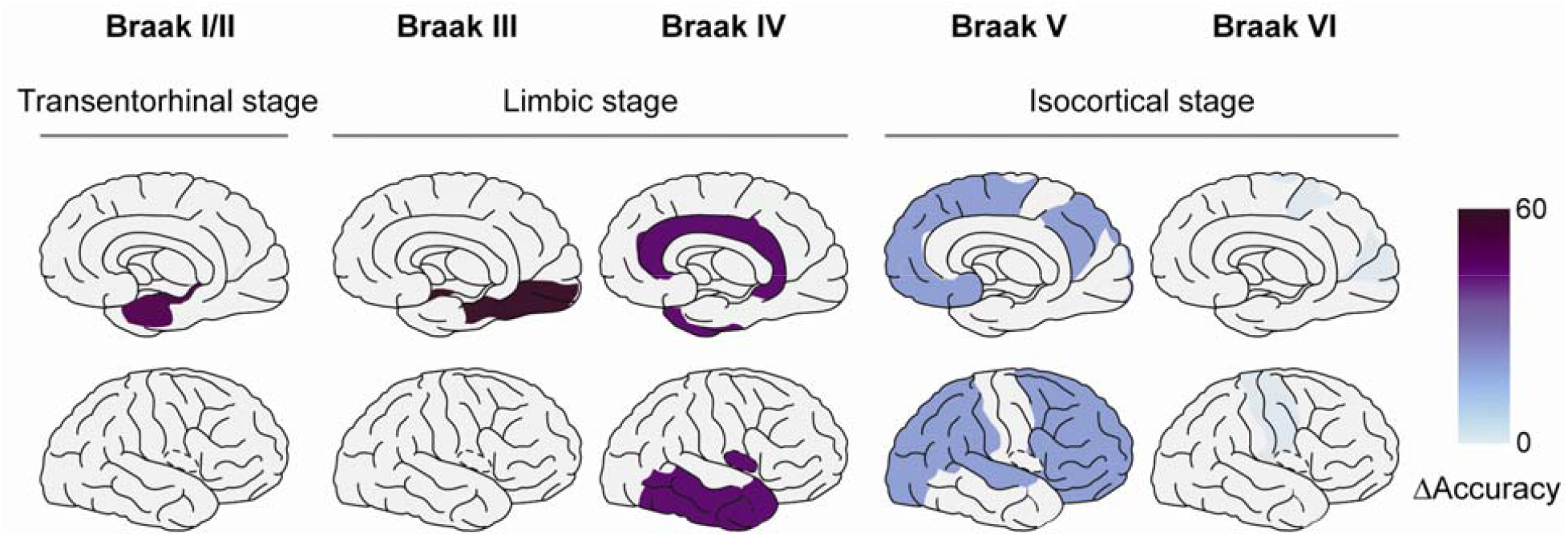
Occlusion of Braak staging ROIs. The results of the occlusion analysis are shown where composite ROIs corresponding to 5 Braak stages (stages I/II, III, IV, V, and VI) were occluded, evaluating their contribution to differentiating between pMCI and sMCI. The Δ*Acc* of each composite Braak staging ROI superimposed on a medial and radial cortical surface model. Abbreviations: pMCI=progressive mild cognitive impairment, sMCI=stable mild cognitive impairment, ROI=region of interest.

## Discussion

While progressive brain atrophy is a ubiquitous consequence of AD neurodegeneration, the major spatial patterns of atrophy observed when individuals progress from the prodromal (MCI) to the clinical phases of AD are not understood well. The motivation of the current study was to identify the major spatial patterns of atrophy predicative of AD progression by leveraging an interpretable deep learning modeling approach. To that end, we propose a framework based on a prognostic convolutional neural network model and occlusion analysis to investigate the regional contribution of atrophy patterns to the prediction of AD progression. Our use of occlusion analysis, in particular, yielded explainable results wherein the contribution of specific brain regions to the performance of the model, and by proxy to AD progression can be evaluated and visualized while holding the contribution of (atrophy in) other regions constant. We found that the hippocampus, fusiform and inferior temporal gyri and precuneus were the strongest contributors to the prediction of AD progression. When considering the medial temporal lobe separately, our analysis revealed larger effects after hippocampal and amygdala occlusion. Finally, we validated our approach by occluding ROIs motivated by the influential Braak staging scheme (Braak & Braak, 1991), finding that the occlusion of a stage III (limbic) ROI had the largest impact on the performance of the model.

We report that atrophy in bilateral hippocampus, fusiform and inferior temporal gyri, and precuneus played a more central role than other regions in the prediction of progression from MCI to AD. These findings are consistent with studies reporting that hippocampus, fusiform, and inferior temporal gyri are affected in early stages of AD (Convit et al., 1997; Jack et al., 1999a; De Santi et al., 2001). Additionally, we evaluated the spatial precision of our method by performing patch-based occlusion analysis focused more locally on the medial temporal lobe. In line with previous studies (Convit et al., 2000; Grundman et al., 2002; Mitolo et al., 2019) voxels in the hippocampus and amygdala contributed the most to the performance of the model, and by proxy to AD progression. Combined, our findings demonstrate that patterns of cortical atrophy, particularly in medial temporal lobe regions, differentiate individuals with MCI into those more and less likely to progress to AD. These findings may also reflect heightened levels of structural and functional redundancy in these regions, which may offer protection against cognitive decline during aging (Langella, Mucha, et al., 2021; Sadiq et al., 2021; Langella, Sadiq, et al., 2021).

We validated our approach of combining a prognostic deep learning model with occlusion analysis, by occluding ROIs based on the Braak staging scheme (Braak & Braak, 1991), and evaluating whether this step resulted in stage-appropriate results. According to this influential staging scheme neurofibrillary pathology in AD starts in transentorhinal cortex, spreading into entorhinal cortex, hippocampus, amygdala and inferior temporal cortex and eventually to other neocortical regions (Braak & Braak, 1991). Studies utilizing MRI have demonstrated that the trajectory of atrophy changes in AD is largely consistent with Braak’s staging scheme (Jack et al., 2002; Silbert et al., 2003; Burton et al., 2009; Whitwell et al., 2012). In the current study we found that occlusion of ROIs corresponding to Braak stage III had a larger effect on the performance of the prognostic model, relative to occlusion of Braak I/II ROIs. These results highlight the importance of limbic areas in the progression from MCI to AD. Moreover, the effect of occlusion gradually decreased when ROIs from later Braak stages were occluded. These findings are consistent with previous studies, documenting brain atrophy in Braak III regions during the prodromal stages of AD (Desikan et al., 2008; Yao et al., 2012), as well as with the early involvement of entorhinal cortex and hippocampus along the AD continuum (Jack et al., 1999b; Devanand et al., 2007). Altogether, our findings suggest that patterns of atrophy starting from the entorhinal cortex and accumulating up to Braak III regions differentiated subjects with progressive MCI from those with a more stable MCI diagnosis, consistent with the former group being composed of subjects who can be viewed as being at the prodromal stage of AD.

Deep learning models have been applied in multiple biomedical image analysis tasks, including segmentation (Chen et al., 2018; Kamnitsas et al., 2017), reconstruction (Schlemper et al., 2018; G. Yang et al., 2018), and classification (Suk et al., 2014; Hosseini-Asl et al., 2016; Cheng et al., 2019). Emphasis on interpretability has increased in recent years, aimed at better understating how and why extracted features contribute to successful class prediction. However, studies have shown that methods such as class activation maps often fail to provide sufficient interpretable findings in models based on medical imaging data (Reyes et al., 2020; Saporta et al., 2021). On the other hand, occlusion analysis, as has been applied here, has been successfully applied as an interpretable approach in similar settings (Kermany et al., 2018; Lu et al., 2021). We suggest that, when combined with a diagnostic or prognostic machine learning model, occlusion analysis can serve as a useful approach for studying the contribution of complex image-based features to various disease states.

Several limitations of our study should be acknowledged. First, although we used one of the biggest publicly-available AD datasets, we acknowledge that future work could benefit from working with larger sample sizes. Second, our model is based solely on structural MRI data. Incorporating other imaging modalities or fluid biomarker data, to assess the contribution of amyloid and tau burden and their synergies with atrophy (Sadiq et al., 2022) may further benefit the performance of the model and should be considered in future studies. Third, our study evaluated the major regional patterns of atrophy predicative of subsequent progression from MCI to AD. We acknowledge that incorporating longitudinal neuroimaging into a similar design could provide additional valuable information on the spatiotemporal patterns of atrophy seen in AD progression. This could also be achieved in future research.

## Conclusion

In conclusion, we identified the major regional substrates of cortical atrophy, predictive of AD progression using a combination of deep learning and occlusion. In agreement with previous results, we found that atrophy in the hippocampus, fusiform, and inferior temporal gyri, were the strongest predictors of AD progression. These results further establish the potential for early identification of individuals with MCI who will likely progress to AD based on patterns of cortical atrophy.

## Supporting information

SI for Kwak et al

## Data Availability

All data produced are available online at: http://adni.loni.usc.edu/

## Acknowledgements

Funding: Research reported in this publication was supported by the National Institute On Aging of the National Institutes of Health under Award Number R01AG062590 and by the UNC Idea Grant. The content is solely the responsibility of the authors and does not necessarily represent the official views of the National Institutes of Health. Data collection and sharing for this project was funded by the Alzheimer’s Disease Neuroimaging Initiative (ADNI) (National Institutes of Health Grant U01 AG024904) and DOD ADNI (Department of Defense award number W81XWH-12-2-0012). ADNI is funded by the National Institute on Aging, the National Institute of Biomedical Imaging and Bioengineering, and through generous contributions from the following: AbbVie, Alzheimer’s Association; Alzheimer’s Drug Discovery Foundation; Araclon Biotech; BioClinica, Inc.; Biogen; Bristol-Myers Squibb Company; CereSpir, Inc.; Cogstate; Eisai Inc.; Elan Pharmaceuticals, Inc.; Eli Lilly and Company; EuroImmun; F. Hoffmann-La Roche Ltd and its affiliated company Genentech, Inc.; Fujirebio; GE Healthcare; IXICO Ltd.;Janssen Alzheimer Immunotherapy Research & Development, LLC.; Johnson & Johnson Pharmaceutical Research & Development LLC.; Lumosity; Lundbeck; Merck & Co., Inc.;Meso Scale Diagnostics, LLC.; NeuroRx Research; Neurotrack Technologies; Novartis Pharmaceuticals Corporation; Pfizer Inc.; Piramal Imaging; Servier; Takeda Pharmaceutical Company; and Transition Therapeutics. The Canadian Institutes of Health Research is providing funds to support ADNI clinical sites in Canada. Private sector contributions are facilitated by the Foundation for the National Institutes of Health (www.fnih.org). The grantee organization is the Northern California Institute for Research and Education, and the study is coordinated by the Alzheimer’s Therapeutic Research Institute at the University of Southern California. ADNI data are disseminated by the Laboratory for Neuro Imaging at the University of Southern California.

## Author contribution

K.K and E.D conceived research; K.K analyzed data; K.K, W.S, and E.D interpreted results and wrote the paper.

