## Supplementary material for "Identifying the regional substrates predictive of Alzheimer’s disease progression through a convolutional neural network model and occlusion": SI for Kwak et al

Eran Dayan, Ph.D.

Address: 130 Mason Farm Road, CB 7513, Chapel Hill, NC, 27599

^†^Data used in the preparation of this article were obtained from the Alzheimer's Disease Neuroimaging Initiative (ADNI) database (<http://adni.loni.usc.edu>). As such, the investigators within the ADNI contributed to the design and implementation of the ADNI and/or provided data but did not participate in analysis or writing of this article. A complete listing of ADNI investigators can be found at <http://adni.loni.usc.edu/wp-content/uploads/how_to_apply/ADNI_Acknowledgement_List.pdf>.

**Table S1. Composite ROIs corresponding to Braak stages.**

| Stage | Regions |
| --- | --- |
| Braak I/II | Entorhinal, Hippocampus |
| Braak III | Parahippocampal, fusiform, lingual, amygdala |
| Braak IV | Middle temporal, Caudate anterior cingulate, Rostral anterior cingulate, Posterior cingulate, Isthmus cingulate, insula, inferior temporal, Temporal pole |
| Braak V | Superior frontal, Lateral orbito frontal, Medial orbito frontal, Frontal pole, Caudal middle frontal, Rostral middle frontal, Pars opercularis, Pars orbitalis, Pars triangularis, Lateral occipital, Parietal supramarginal, Parietal inferior, Superior temporal, Parietal superior, Precuneus, Bank Superior Temporal, Transverse temporal |
| Braak VI | Pericalcarine, Postcentral, Cuneus, Precentral, Paracentral |

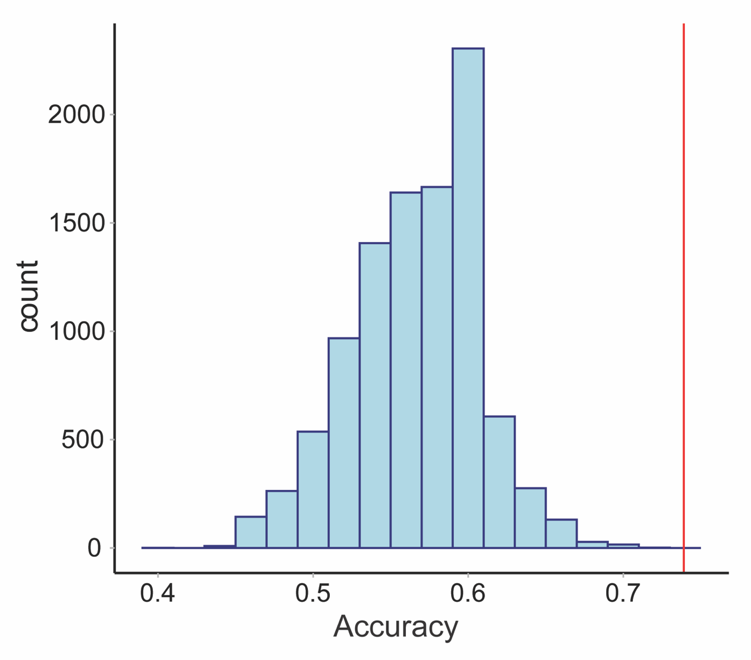

**Figure S1. Permutation scores (null distribution) obtained by a random classifier model.** Null distribution of accuracy levels generated with a random classifier deployed on 10000 different permutations of the dataset. The red line indicates the accuracy obtained by the proposed model using the original data.

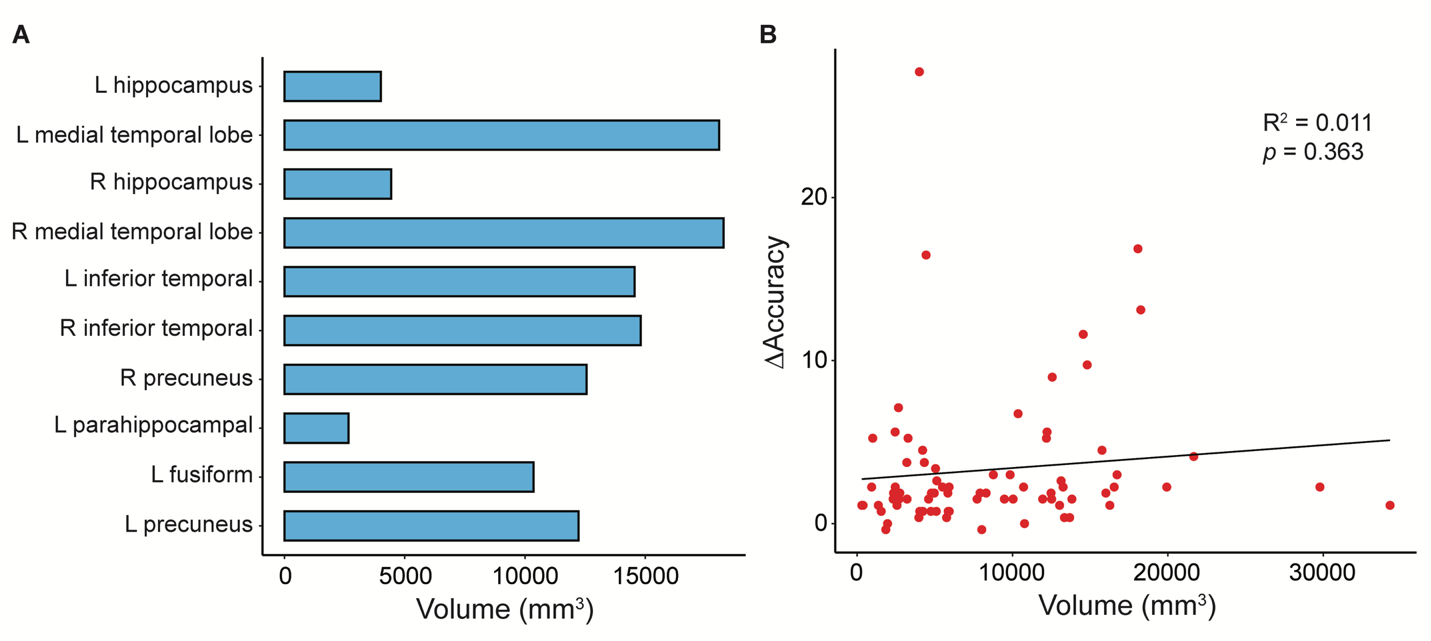

**Figure S2. The effect of regional volume size on the results of the occlusion analysis.** (**A**) Volume size is shown for the top 10 regions in descending order according to the results of the occlusion analysis. (**B**) The relationship between delta accuracy and regional volume size was not statistically significant.
